# OpenSAFELY: Representativeness of Electronic Health Record platform OpenSAFELY-TPP data compared to the population of England

**DOI:** 10.1101/2022.06.23.22276802

**Authors:** Colm D Andrews, Anna Schultze, Helen J Curtis, William J Hulme, John Tazare, Stephen JW Evans, Amir Mehrkhar, Seb Bacon, George Hickman, Chris Bates, John Parry, Frank Hester, Sam Harper, Jonathan Cockburn, David Evans, Tom Ward, Simon Davey, Peter Inglesby, Ben Goldacre, Brian MacKenna, Laurie Tomlinson, Alex J Walker

**Affiliations:** The DataLab, Nuffield Department of Primary Care Health Sciences, University of Oxford, OX26GG; London School of Hygiene and Tropical Medicine, Keppel Street, London WC1E 7HT; TPP, TPP House, 129 Low Lane, Horsforth, Leeds, LS18 5PX

**Keywords:** OpenSafely, Representative

## Abstract

**Background:** Since its inception in March 2020, data from the OpenSAFELY-TPP electronic health record platform has been used for more than 50 studies relating to the global COVID-19 emergency. OpenSAFELY-TPP data is derived from practices in England using SystmOne software, and has been used for the majority of these studies. We set out to investigate the representativeness of OpenSAFELY-TPP data by comparing it to national population estimates.

**Methods:** With the approval of NHS England, we describe the age, sex, Index of Multiple Deprivation and ethnicity of the OpenSAFELY-TPP population compared to national estimates from the Office for National Statistics. The five leading causes of death occurring between the 1st January 2020 and the 31st December 2020 were also compared to deaths registered in England during the same period.

**Results:** Despite regional variations, TPP is largely representative of the general population of England in terms of IMD (all within 1.1 percentage points), age, sex (within 0.1 percentage points), ethnicity and causes of death. The proportion of the five leading causes of death is broadly similar to those reported by ONS (all within 1 percentage point).

**Conclusions:** Data made available via OpenSAFELY-TPP is broadly representative of the English population.

**Summary:** Users of OpenSAFELY must consider the issues of representativeness, generalisability and external validity associated with using TPP data for health research. Although the coverage of TPP practices varies regionally across England, TPP registered patients are generally representative of the English population as a whole in terms of key demographic characteristics.

**Key messages:**

- There is regional variability across England in terms of key population characteristics
- Users of OpenSAFELY should carefully consider the issues of representativeness, generalisability and external validity associated with using TPP data for health research.
- TPP registered patients are a representative sub-sample of the English population as a whole in terms of age, sex, IMD and ethnicity.
- The proportions of the five leading causes of death in TPP in 2020 are broadly similar to those reported by ONS.

## Background

OpenSAFELY is a secure health analytics platform created by our team on behalf of NHS England. OpenSAFELY provides a secure software interface allowing analysis of pseudonymised primary care patient records from England in near real-time within highly secure data environments. OpenSAFELY software is currently deployed within the secure data centres of the two largest electronic health record providers in the NHS: EMIS and TPP, and is delivering federated analytics where the same data curation and analysis code executes in each environment. To date more than 20 publications have used the OpenSAFELY platform, focused on delivering vital and urgent results related to the global COVID-19 emergency. Some of these papers have used the federated analytics functionality more latterly available in OpenSAFELY to deliver combined analyses across 58 million patients’ data in both OpenSAFELY-EMIS and OpenSAFELY-TPP (1–5); however the majority of analyses published during the pandemic specifically used OpenSAFELY-TPP which covers 40% of general practices in England, those using SystmOne Electronic Health Record (EHR) software produced by TPP.

The use of data from EHR providers is an invaluable tool for health research, however data is primarily collected for clinical use and not specifically with research in mind. As these datasets are not a random sample of the population of interest, it is important to understand the representativeness of the data. The deployment of TPP SystmOne software is known to be geographically clustered (6), and factors such as sex, age, ethnicity and levels of deprivation, which are important clinical risk factors for death from COVID-19 (7), show regional variability across England (8). Key outcomes such as causes of death also vary by region (9). However, little is currently known about how the characteristics of patients in TPP practices compare to the population at large.

In order to aid the interpretation of ongoing COVID-19 research projects in OpenSAFELY-TPP we therefore set out to compare key demographic characteristics of patients registered with TPP practices to national estimates from the Office for National Statistics (ONS). We also compared the distribution of the five leading causes of death registered in ONS between 1st January 2020 and 31st December 2020 to deaths registered in TPP during the same period.

## Methods

### Study design

#### Data Source

Primary care records managed by the GP software provider TPP were linked to ONS death data through OpenSAFELY-TPP, a data analytics platform created by our team on behalf of NHS England to address urgent COVID-19 research questions (https://opensafely.org). Similarly pseudonymized datasets from other data providers are securely provided to the EHR vendor and linked to the primary care data. The dataset analysed within OpenSAFELY-TPP is based on 24 million people currently registered with GP surgeries using TPP SystmOne software. **It includes pseudonymized data such as coded diagnoses, medications and physiological parameters. No free text data are included**. Further details on our information governance can be found on page 20, under <u>information governance and ethics</u>.

The UK Census collects individual and household-level demographic data every 10 years for the whole UK population. Data on ethnicity were obtained from the 2011 UK Census for England. In addition to census data, ONS release annual mid-year estimates of the resident population of England produced using a cohort component method (10). Data on IMD, Age and sex were obtained from the 2020-mid year estimates and estimates of the 5 most common causes of death in 2020 were obtained from ONS mortality statistics published via NOMIS (11).

#### Study population

For demography and coverage analyses, patients were included in the study if they were registered at an English general practice using a TPP SystmOne clinical information system on 30th June 2020. For analysis of causes of death, patients were included if they were registered with an English general practice using a TPP SystmOne clinical information system on the day of a death registered on ONS between 1st January 2020 and 31st December 2020.

#### Demographic Data

Ethnicity: The primary care recorded ethnicity, supplemented where missing with ethnicity data from the Secondary Uses Service (SUS), was collapsed into the five high-level and 16 detailed census categories of White (White British, White Irish, other White), South Asian (Indian, Pakistani, Bangladeshi, other South Asian), Black (African, Caribbean, other Black), other (Chinese, all others), and mixed (White and Asian, White and African, White and Caribbean, other mixed) with an additional unknown ethnicity category included.

Age: Patients’ age was calculated as of 30th June 2020 and grouped into 5 year bands.

Sex: We used categories “male” and “female”, matching the ONS recorded categories; patients with any other/unknown sex were included as “unknown”.

Deprivation: Deprivation was measured by the Index of Multiple Deprivation (IMD) derived from the patient’s postcode at lower super output area level. IMD was divided into quintiles, with higher values indicating greater deprivation.

#### Causes of Death

Patients were flagged if they had any death certified and registered in England or Wales between 1st January 2020 and 31st December 2020 and where applicable grouped into the 5 most common underlying causes of death (Table 1).

**Table 1:** Most common underlying causes of death occurring in England 2020 (26):

| Cause of Death | ICD 10 codes | N (% of all deaths) |
| --- | --- | --- |
| COVID-19 | U07 | 73680 (13) |
| Dementia and Alzheimer disease | F01-F03, G30 | 70035 (12) |
| Ischaemic heart diseases | I20-I25 | 55690 (10) |
| Cerebrovascular diseases | I60-I69 | 29680 (5) |
| Malignant neoplasm of trachea, bronchus and lung | C33-C34 | 28720 (5) |

### Statistical methods

We investigated the representativeness of TPP data by comparing OpenSAFELY-TPP-derived figures for 2020 with the following: (a) ONS IMD for all of England, (b) ONS age, sex (2020 estimates) and ethnicity (2011 census) across NHS England operating regions, and (c) causes of death (Malignant neoplasm of trachea, bronchus and lung, Ischaemic heart diseases, Dementia and Alzheimer disease, COVID-19 and Cerebrovascular diseases) in 2020 across NHS England operating regions. Proportions of each age group, sex, IMD band, ethnicity and cause of death were calculated and compared to the corresponding ONS data. For mortality analysis the denominator was the total number of deaths in 2020 and the numerator was the number of patients with the relevant ICD10 code (Table 1) as the underlying cause. TPP coverage was calculated as the proportion of TPP registered patients compared to ONS estimated populations within each Nomenclature of Territorial Units for Statistics (NUTS 1) region.

### Software and Reproducibility

Data management was performed using Python 3.8, with analysis carried out using R. Code for data management and analysis as well as codelists are openly available online at (https://github.com/opensafely/representativeness) for inspection and re-use by anyone.

### Patient and Public Involvement

We have developed a publicly available website https://opensafely.org/ through which we invite any patient or member of the public to contact us regarding this study or the broader OpenSAFELY project.

## Results

### TPP coverage

The population of active TPP patients (alive and registered on 30th June 2020) was 24 million representing 42.6% of the total UK population (based on the UK 2020 mid-year population estimate of 56 million). TPP coverage as a proportion of the ONS population was highest in the East of England (90.5%) and East Midlands (86.1%) and lowest in the West Midlands (16.8%), South East England (17.6%) and London (18.7%) (Figure 1).

**Figure 1:**
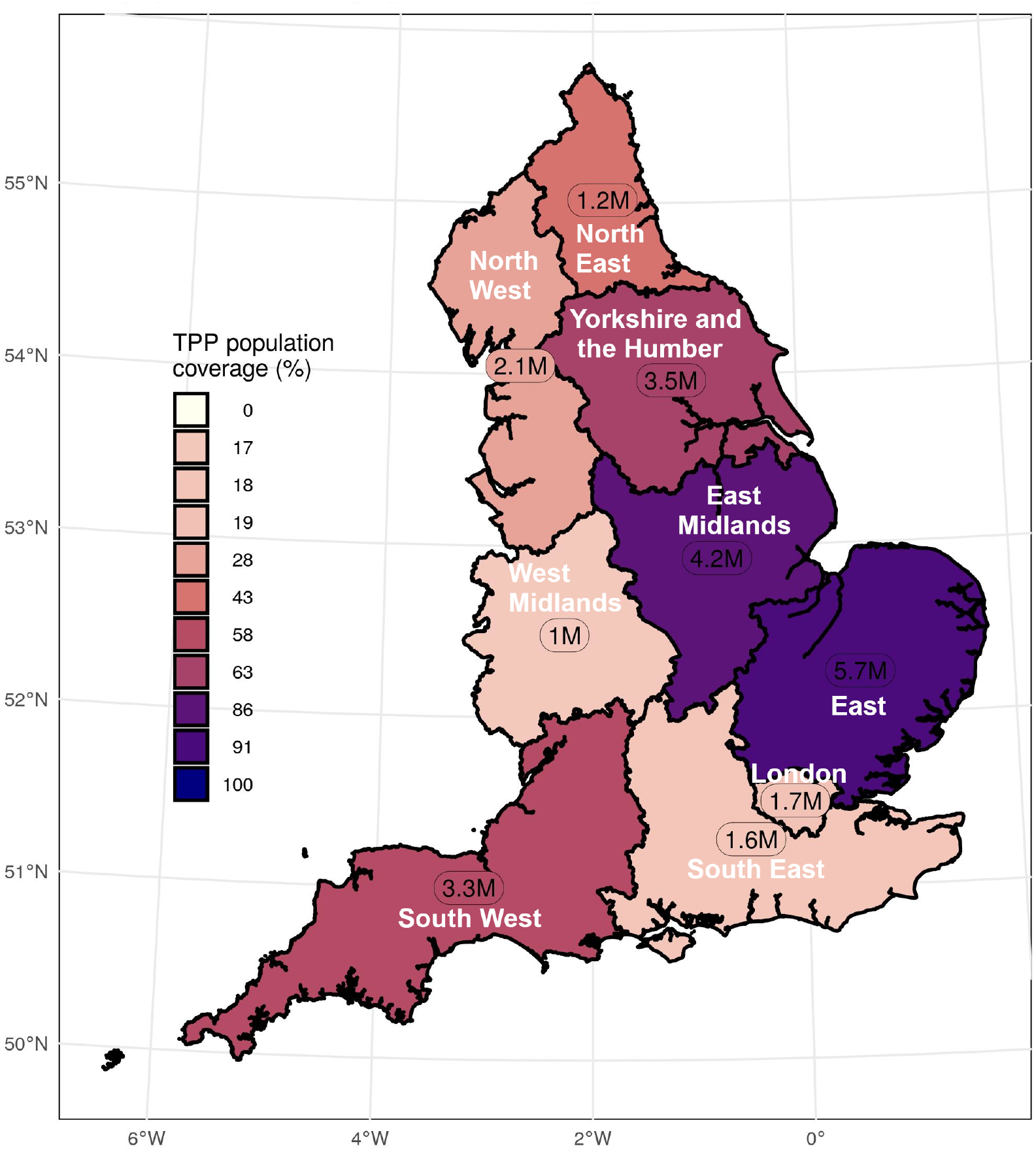
Population coverage map showing coverage of each Nomenclature of Territorial Units for Statistics (NUTS-1) region. Population coverage based on ONS estimates covered by TPP with the number of patients in TPP per region.

### IMD

Overall the proportion of IMD groups was similar, with only small differences between the TPP and ONS populations: In those with a recorded IMD there was a slightly higher proportion of TPP patients in the most deprived IMD group 1 (20.5%) and IMD group 3 (21.1) compared to national ONS estimates (20.0 and 20.3 respectively). TPP practices underrepresented patients in the least deprived IMD group 5 (18.3%) compared to ONS (19.4%) (Figure 2). IMD was missing for 2.3% of the TPP records.

**Figure 2:**
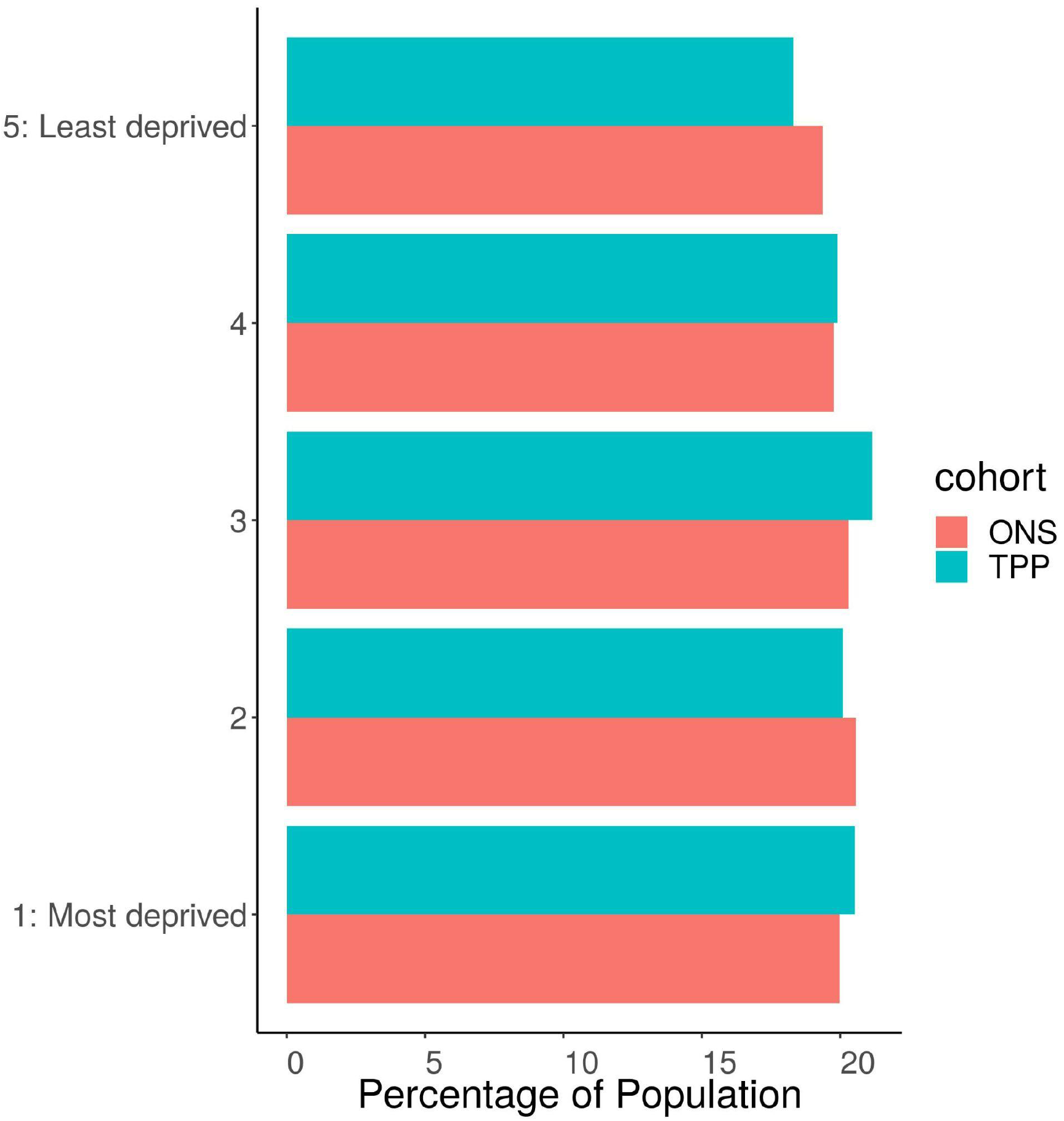
Barplot showing the proportion of ONS and TPP populations per IMD Quintile. The TPP population excludes 2.3% of patients without a recorded IMD.

### Sex

For those with sex recorded as either male or female there was a similar proportion of women in the English population (50%) compared to ONS (50.1%) (Figure 3). The South West of England had the highest proportion of Females in TPP (50.8%) with London having the lowest proportion (48.8%). The difference in proportion of women between TPP and ONS estimates was within 0.1 percentage points for all regions (Figure 4).

**Figure 3:**
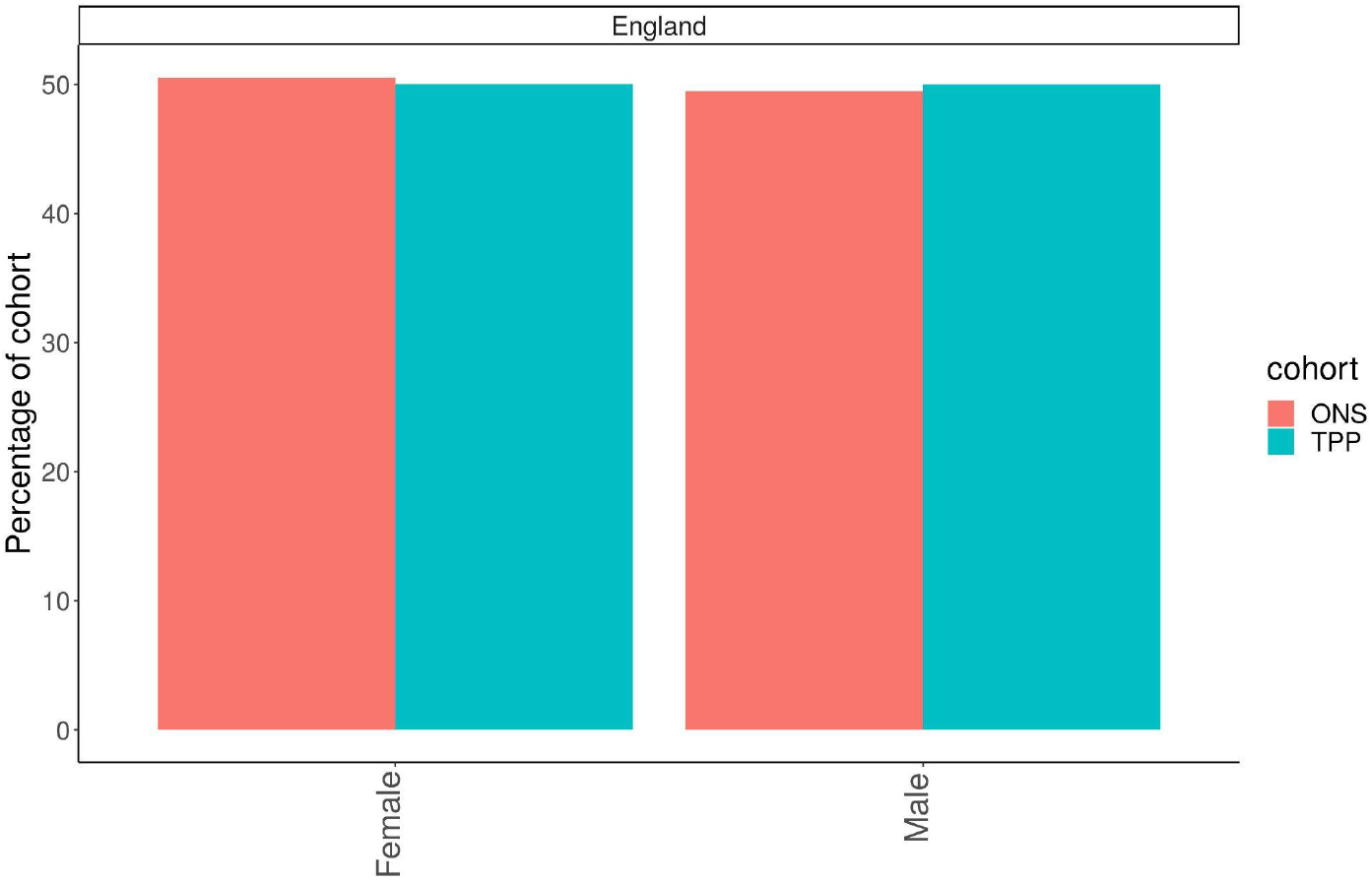
Barplot showing the proportion of ONS and TPP populations by Sex

**Figure 4:**
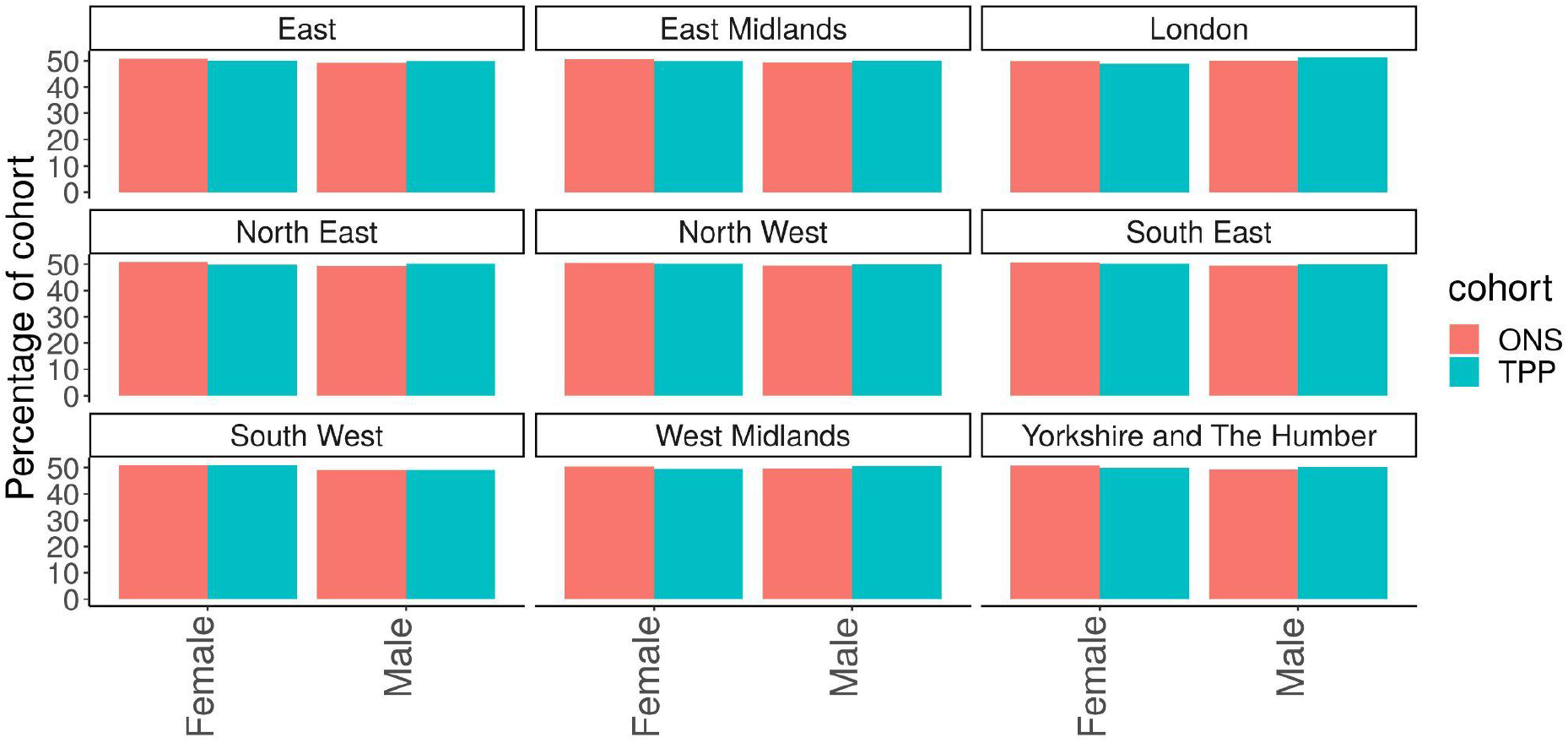
Barplot showing the proportion of ONS and TPP populations by Sex per NUTS-1 region

### Age

There was a higher proportion of TPP patients in the age range 25-59 compared to ONS nationally, with a lower proportion of those under 25 years old (Figure 5). The difference in age distribution between TPP and ONS estimates was highest in London and the age distribution of the South West most closely resembled the ONS estimates (Figure 6).

**Figure 5:**
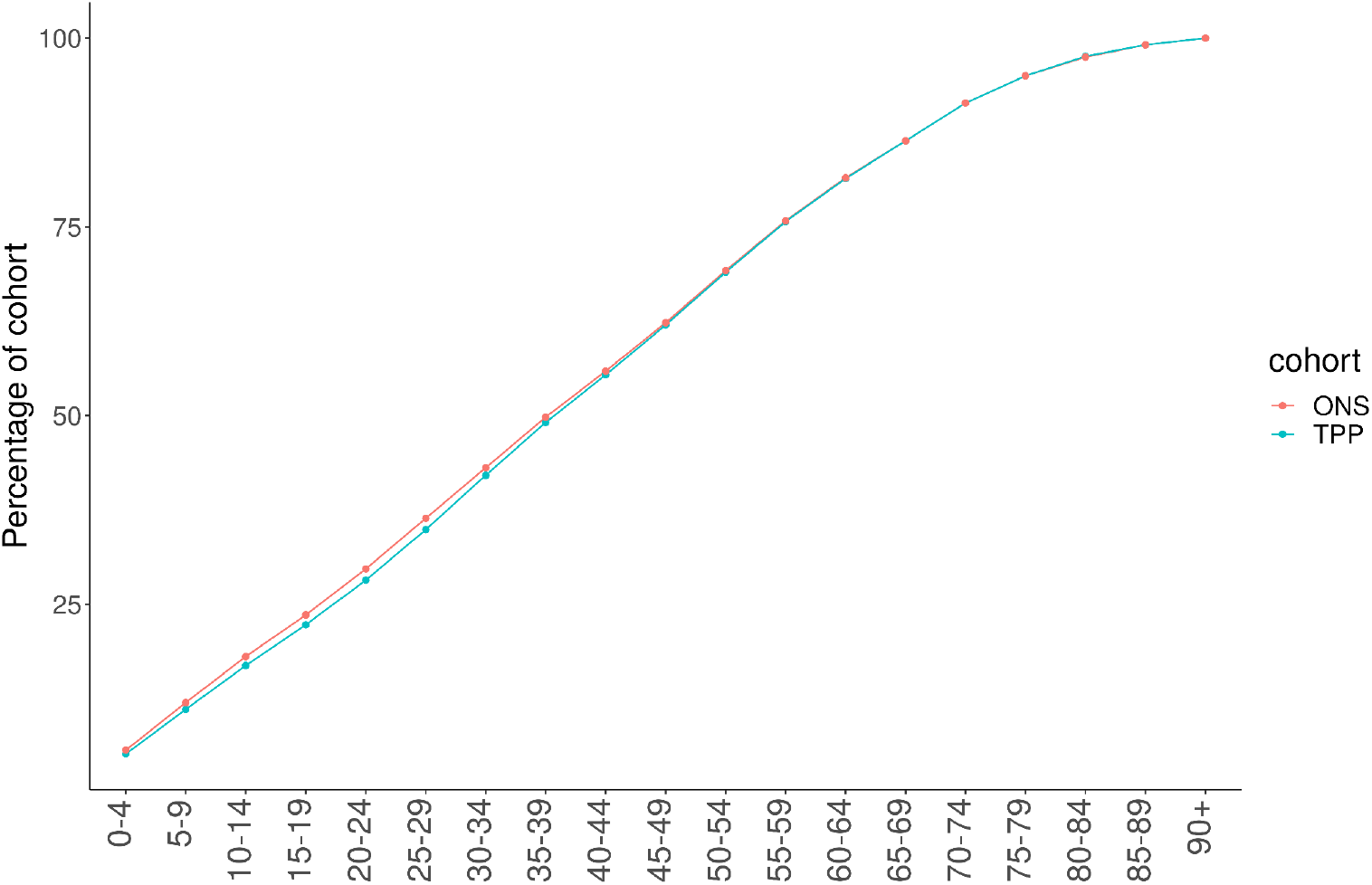
Cumulative frequency graph of ONS and TPP populations by age band

**Figure 6:**
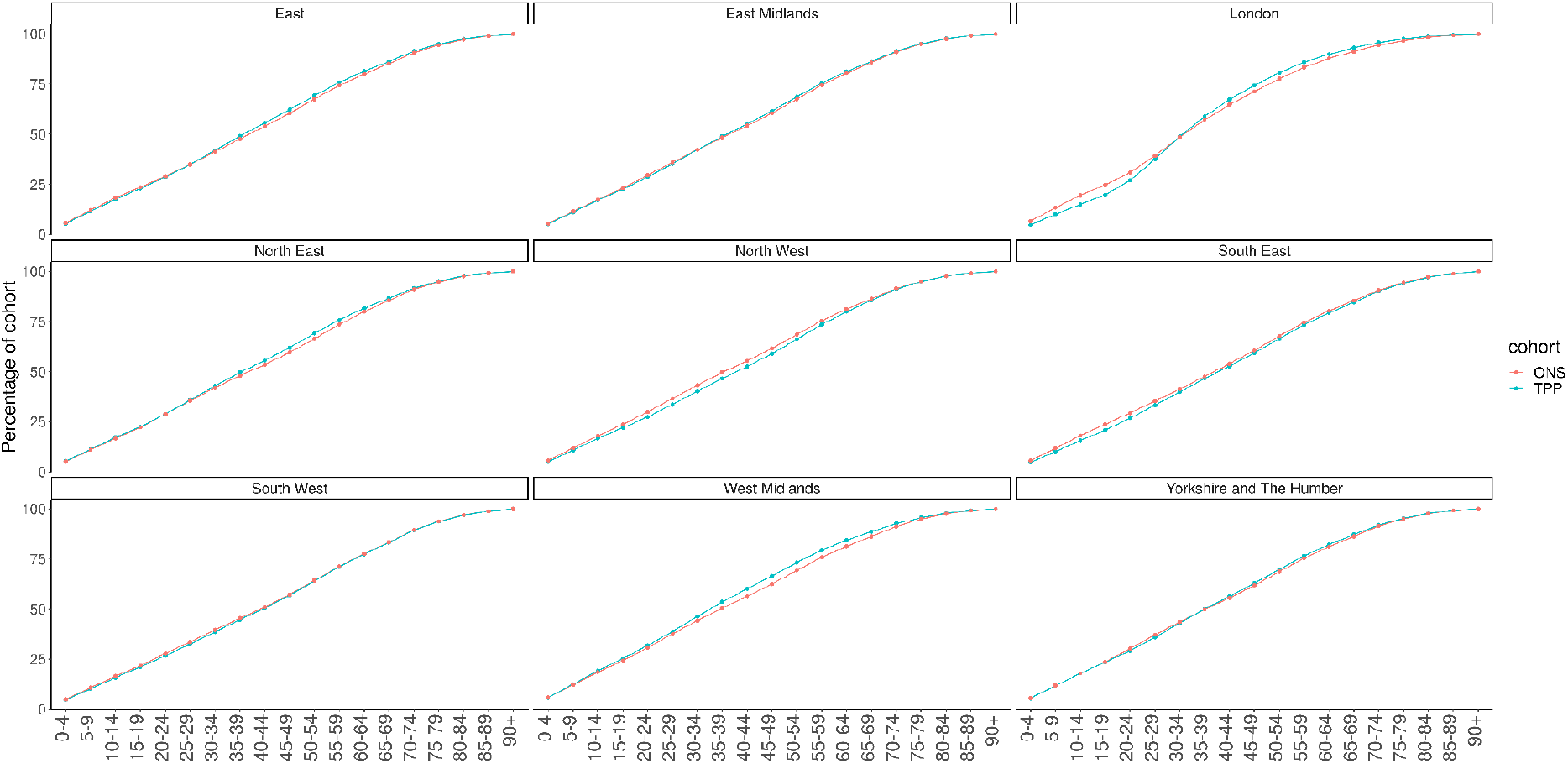
Cumulative frequency graph of ONS and TPP populations by age band per NUTS-1 region

### Age and sex

Across England as a whole the higher proportion of TPP patients in the 35-59 age range compared to ONS estimates was largely due to a higher proportion of men in this age group in TPP. There was a higher proportion of women aged 20-29 in TPP and a lower proportion of men aged 20-29 compared to ONS estimates (Figures 7, 8).

**Figure 7:**
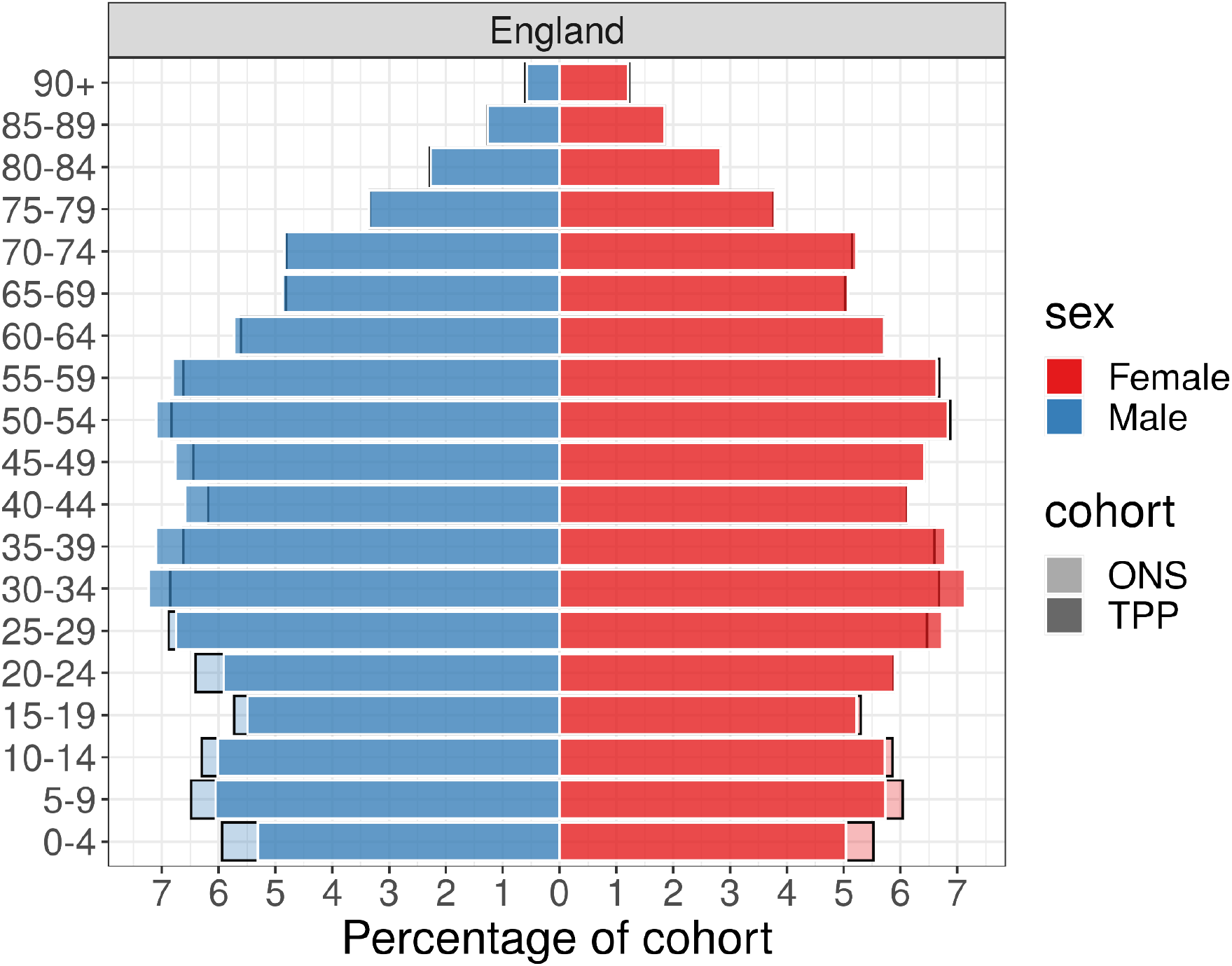
Barplot showing the proportion of ONS and TPP populations by sex and age band

**Figure 8:**
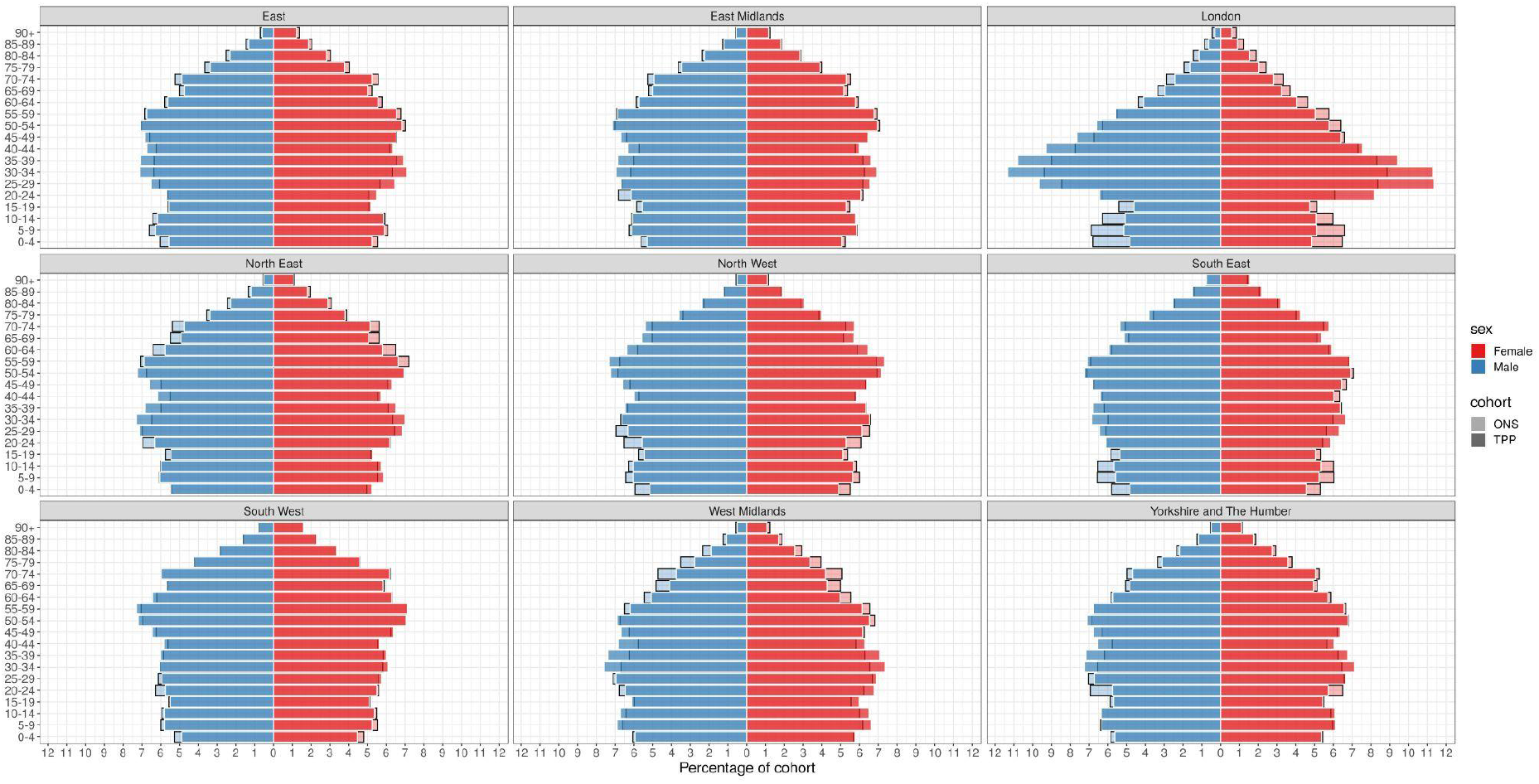
Barplot showing the proportion of ONS and TPP populations by sex and age band per NUTS-1 region

### Causes of death

Across England there was a lower proportion of all five of the leading causes of deaths in TPP compared with ONS data (Figure 9). The biggest difference was in COVID-19 (12.2% in TPP, 12.9% in ONS) and the smallest difference was in Malignant neoplasm of trachea, bronchus and lung (4.9% in TPP, 5.0% in ONS). The difference in proportions of all 5 leading causes of death compared to ONS varied by region (Figure 10). COVID was overrepresented in TPP in all regions other than the North West (14.9% in TPP, 14.9% in ONS) and South East (7.5%, 10.0%).

**Figure 9:**
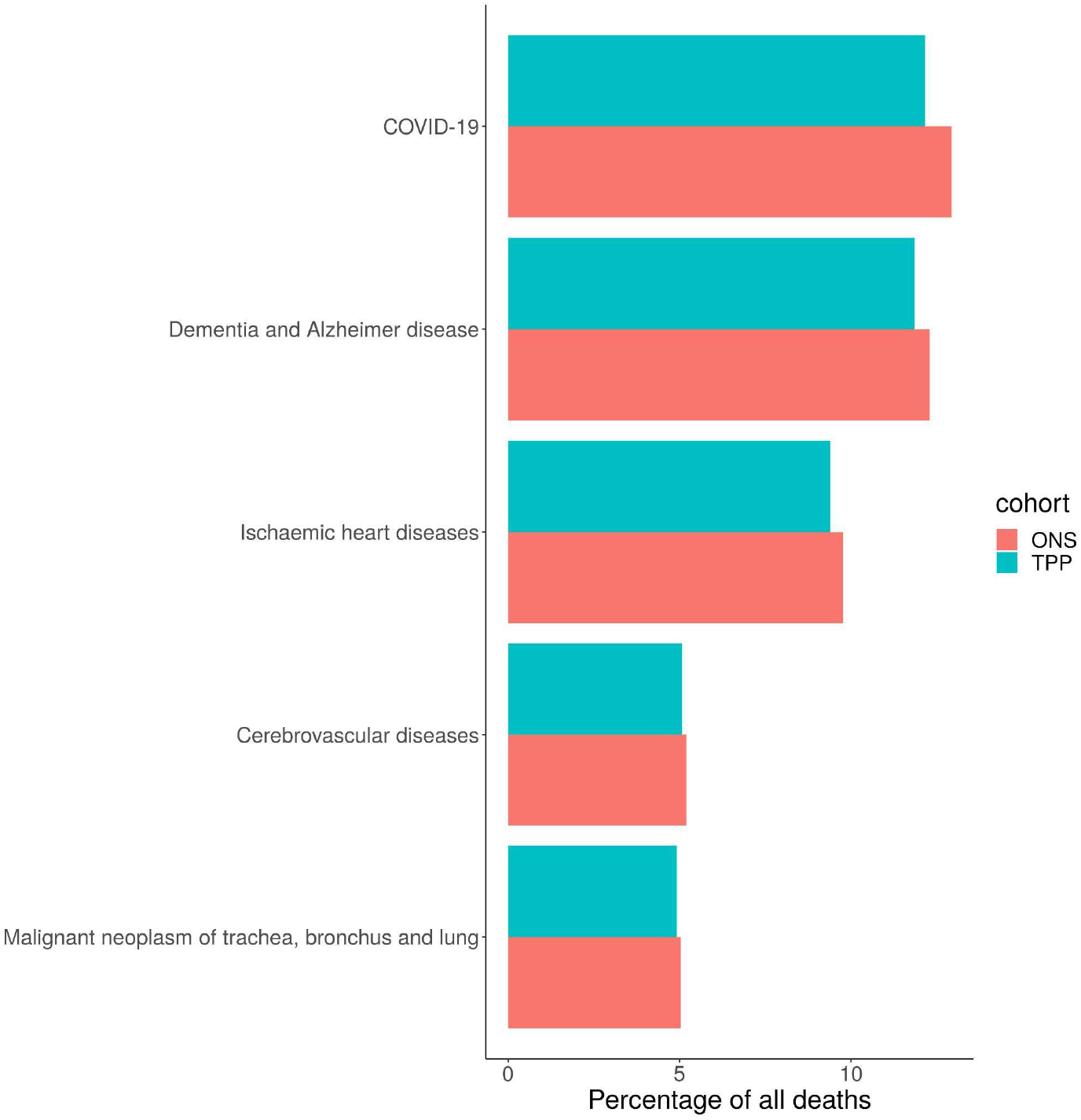
Barplot showing the proportion of the 5 most common causes of deaths occuring in ONS and TPP in 2020.

**Figure 10:**
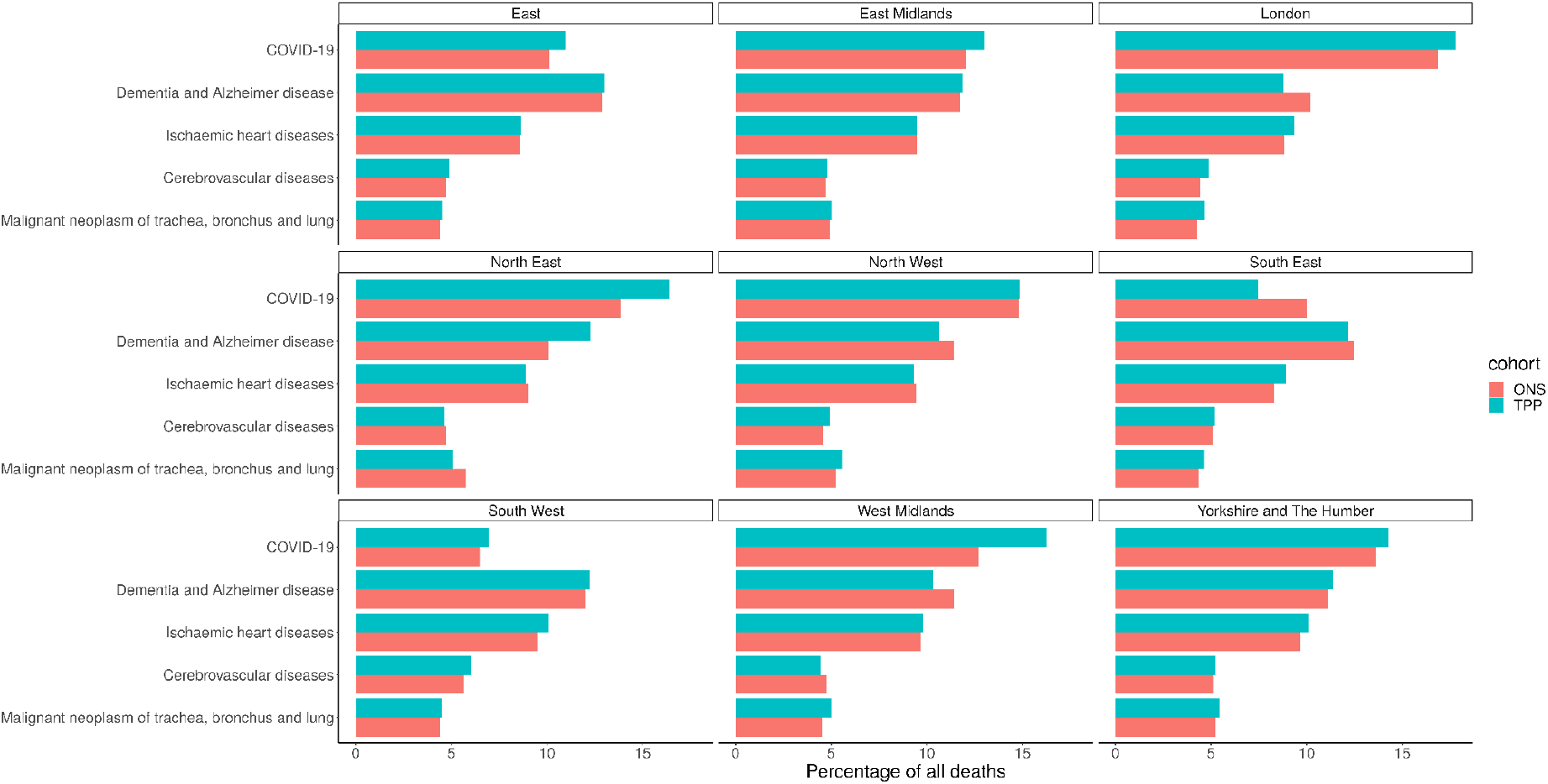
Barplot showing the proportion of the 5 most common causes of deaths occuring in ONS and TPP in 2020 per NUTS-1 region

### Ethnicity (5 Groups)

Of those with a recorded ethnicity the proportion of each ethnic group was within 1 percentage point of the ONS estimate across England as a whole for the 5 group ethnicity (Figure 11A, 12). The White population was underrepresented in all regions other than the North West (93.3%, 90.2%) (Figure 13). The Asian population was overrepresented in all regions other than the North West (3.5%, 5.5%) and South East (3.9%, 4.6%) (Figure 14). Ethnicity was not recorded for 9.4% of the TPP population.

**Figure 11:**
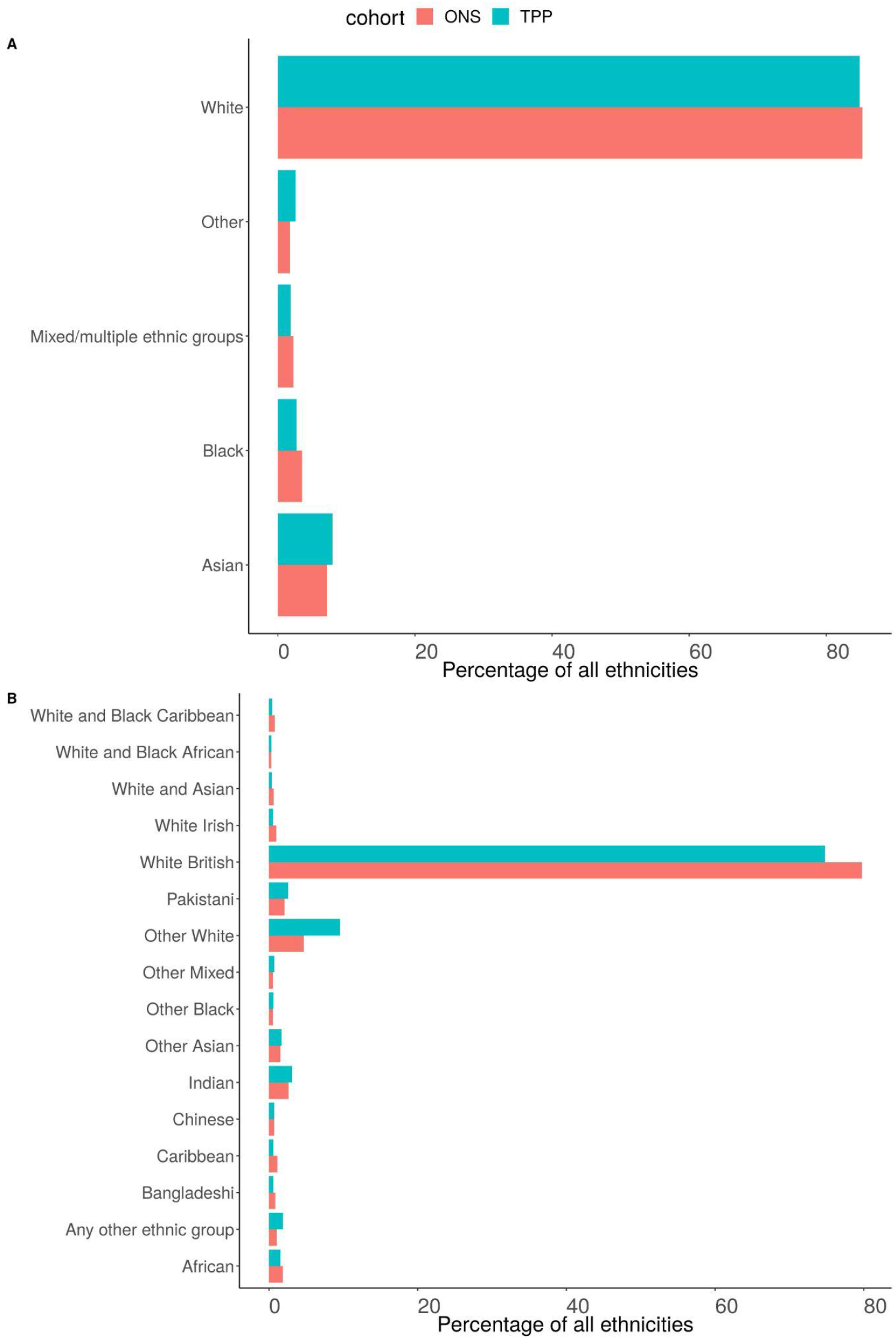
Barplot showing the proportion of ONS and TPP populations per ethnicity grouped into A) 5 and B) 16 groups. The TPP population excludes the 9.4% without a recorded ethnicity.

**Figure 12:**
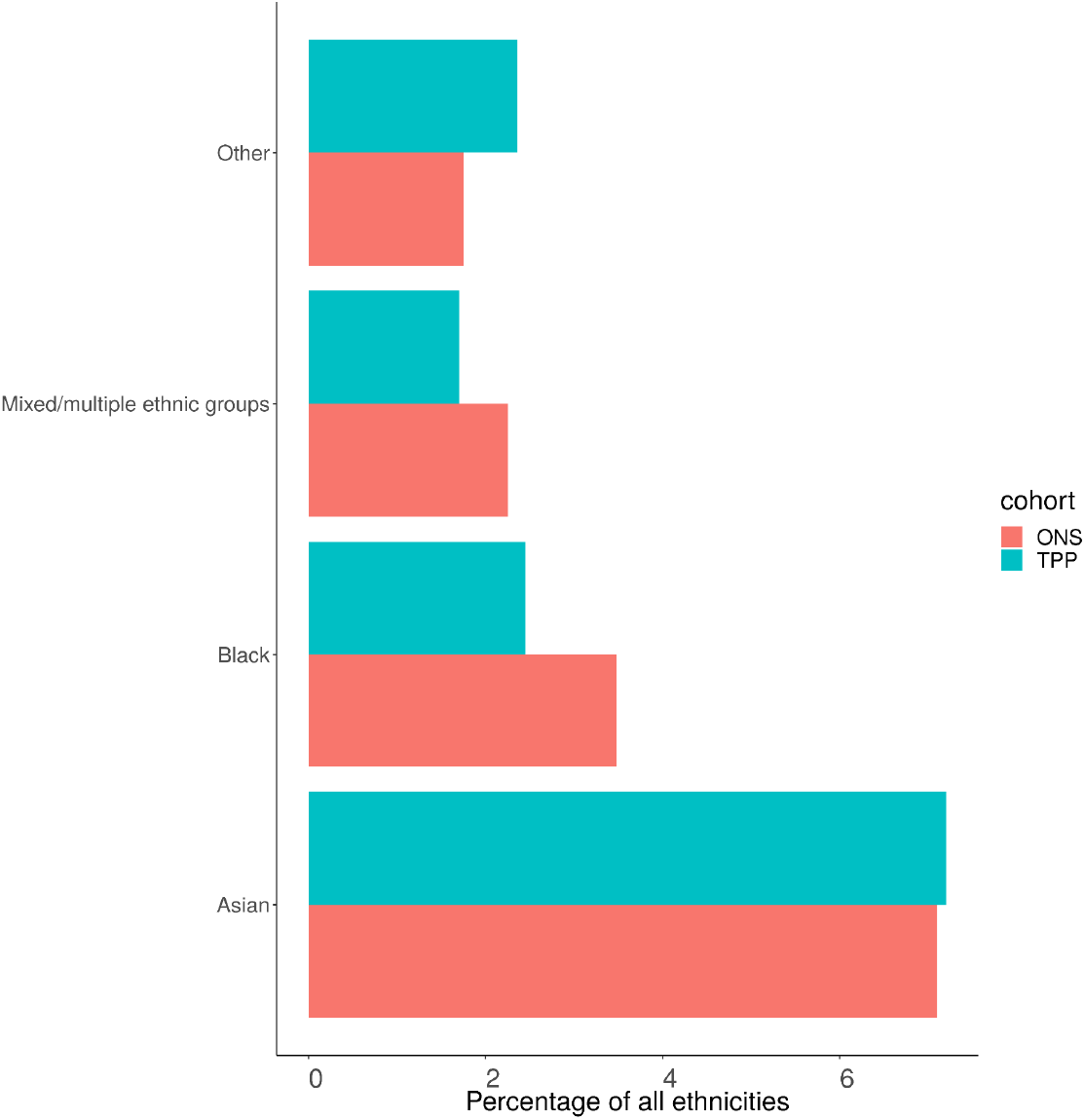
Barplot showing the proportion of ONS and TPP populations per ethnicity (excluding White group)

**Figure 13:**
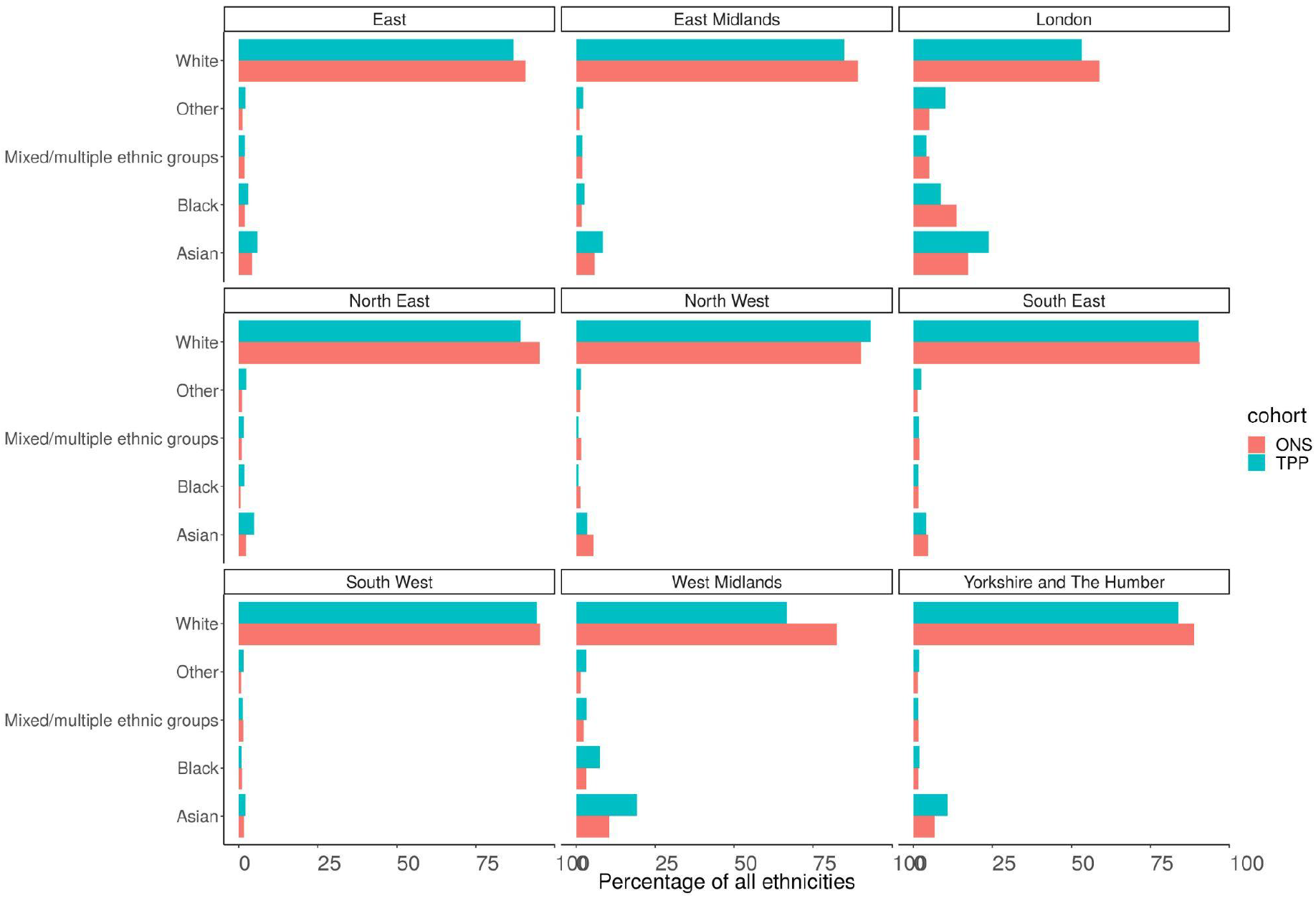
Barplot showing the proportion of ONS and TPP populations per ethnicity grouped into 5 groups per NUTS-1 region

**Figure 14:**
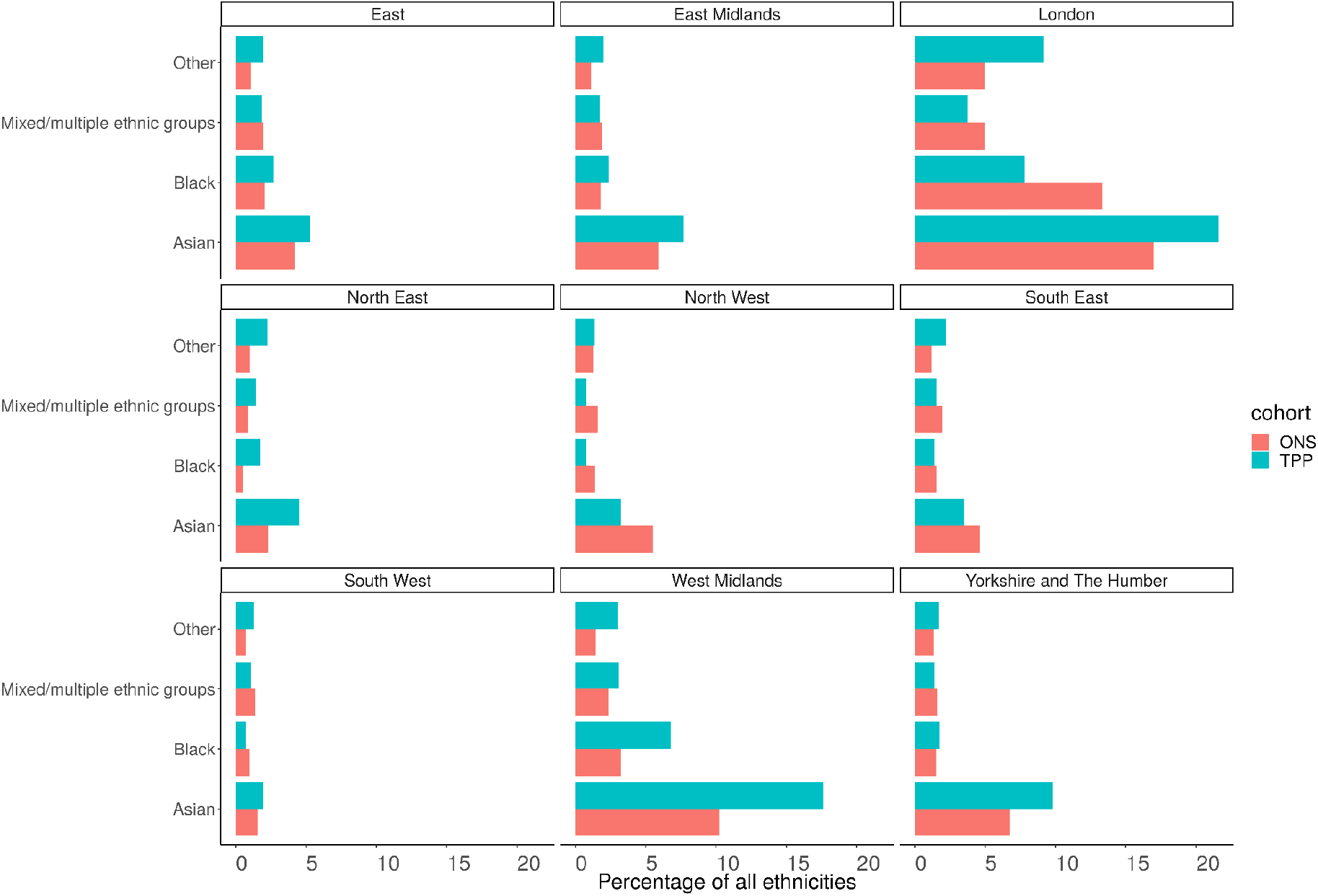
Barplot showing the proportion of ONS and TPP populations per ethnicity grouped into 5 groups per NUTS-1 region (excluding White group)

### Ethnicity (16 groups)

Of those with a recorded ethnicity there was a lower proportion of White British people in TPP (74.8%) compared to ONS (79.8%) and higher proportion of Other White patients (9.6% TPP, 4.7% ONS). There was a lower proportion of both African (1.5%, 1.8%) and Caribbean (0.6%, 1.1%) patients and a higher proportion of Other black patients (0.6%,0.5%) (Figure 11B). There was clear regional variation in both the ethnic makeup of populations and the representativeness of ethnicity in TPP (Figure 15).

**Figure 15:**
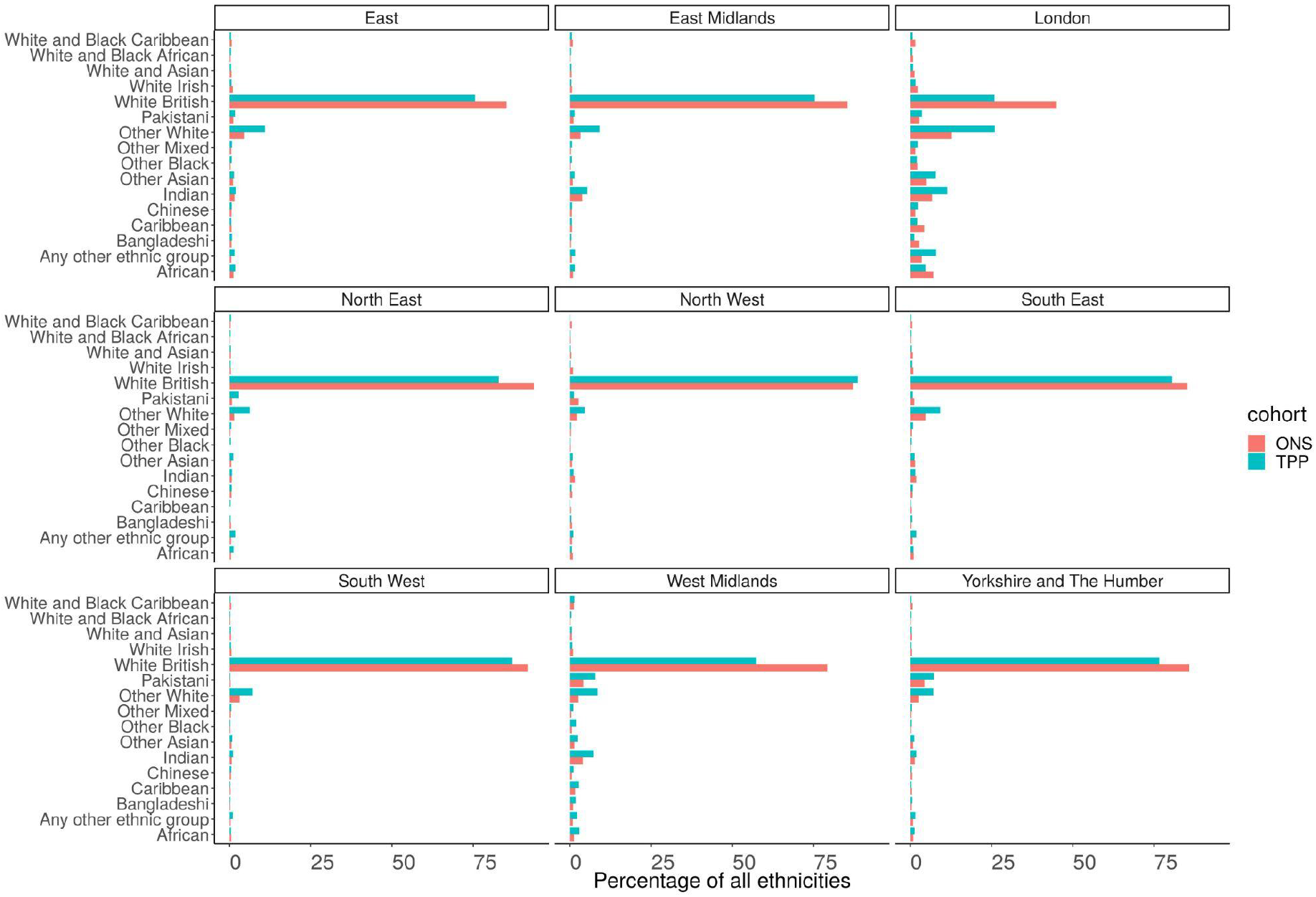
Barplot showing the proportion of ONS and TPP populations per ethnicity grouped into 16 groups per NUTS-1 region

## Discussion

### Summary

This study has shown that TPP data made available via OpenSAFELY-TPP is broadly representative of the English population. Though there is high regional variability in the coverage of the OpenSAFEY-TPP data amongst English general practices, we nonetheless found broad similarity within regions, with only occasional discrepancies which should be considered when designing studies and interpreting outcomes from OpenSAFELY-TPP. Particularly notable was the over-representation of 25-50 year olds in London for both males and females. This may have contributed to a slight overall under-representation of under 25 year olds and over-representation of 25 -59 year olds nationally. We await the 2021 Census results as the assumption that ONS mid-year estimates nearly 10 years after the Census are more accurate than the TPP data may not be true.

### Strengths and weaknesses

This study provides an overview of the representativeness of the OpenSAFELY-TPP cohort with regard to a variety of key characteristics in comparison with the general UK. The key strengths of this study are the use of high quality data from the EHR of all patients registered with a TPP practice which enabled us to compare participation rates for key sociodemographic characteristics (age, sex, IMD, and geographic location) and the comparison of ONS data held in OpenSAFELY-TPP to the matching national ONS data. The true population of England is not known (12), so we compared the OpenSAFELY-TPP data to ONS estimates as it forms the official population estimates of the UK (13). However, ONS estimates may over- or under-estimate population sizes based on regional differences. The mid-year population estimates also have limitations; they heavily rely on the use of health service data to estimate internal migration, in a process that may not be detecting all aspects of population change (12,13). The total GP registered population is always higher than the ONS population nationally, possibly due to over-counting in GP practice registers; under-counting in population estimates and different definitions of who counts as ‘resident’ in the country, but can be lower in certain regions (12). We have, therefore, probably overestimated the overall percentage of the total UK population covered by TPP (42.6%). Compared to the total GP registered population (60 million)(14), TPP covers 39.9% of the UK population. If we considered smaller geographic regions it would be possible to get areas with over 100% coverage.

OpenSAFELY-TPP may have included deaths that were registered in Wales for patients registered in an English practice using TPP, whereas the ONS data was restricted to deaths registered in England.

The most up-to-date formal estimates of the population by ethnic group currently available are from the 2011 Census. The ethnic makeup may have changed substantially from this point. OpenSAFELY-TPP was missing ethnicity for 10% of patients, and the missingness of ethnicity data in EHRs may not be random (8). The 2011 census used multiple imputation to account for missing ethnicity (15).

We investigated the top 5 causes of death (accounting for ∼45%of all deaths) but did not look at others, or at health status more generally e.g. number of long term conditions. Regions are very large and more detailed regional analysis would be informative. We looked at one point in time and representativeness could change with time (e.g. TPP may take on or lose practices) or vary from year to year (e.g. the vaccine campaign may have prompted duplicated patients to be identified and deregistered).

### Findings in Context

Over 50 studies have been conducted using the OpenSafely framework. However, the sheer scale of data made available in OpenSAFELY-TPP alone does not guarantee that the findings are generalisable to the English population at large. While at least one study has shown the large degree of geographical variation in coverage between EHR software providers (16), to our knowledge this is the first time that the representativeness of the population covered by the EHR software provider TPP has been systematically reported. We found that patients in TPP practices are broadly representative of England in terms of age, sex, IMD and ethnicity. The proportion of the five leading causes of death was broadly similar to those reported by ONS. The importance of a representative sample depends on the study question (17,18): careful consideration of this issue is warranted at the design, analysis and interpretation stage of every epidemiological study (19). We have previously described how differences in the design of EHR user interfaces can affect clinical coding (3) and the prescribing of certain medications (20,21). Additionally investigators may wish to consider representativeness of TPP when assessing variation in delivery of healthcare services due to variation in NHS service delivery and TPPs geographical coverage.

### Policy Implications and Interpretation

The breadth of coverage provided by OpenSAFELY-TPP, 24 million patients across England representing 43% of the total English population (based on the ONS 2020 mid-year population estimate of 56 million), provides an unprecedented opportunity to support urgent research into the COVID-19 emergency. Users of OpenSAFELY must consider the issues of representativeness, generalisability and external validity associated with using TPP data for health research: overall, as this analysis shows, TPP registered patients are a representative sample of the English population as a whole.

This paper is principally to inform interpretation of the numerous analyses completed and published using OpenSAFELY-TPP. However, OpenSAFELY is now also implemented in the data analysis environment of EMIS, a primary care electronic health record system supplier covering 55% of all practices in England. In addition, OpenSAFELY also supports federated analytics, where the same data preparation and analysis code is sent to OpenSAFELY-TPP and OpenSAFELY-EMIS to execute the same curation and analysis in each setting, with the results then combined into a single analysis, with a variety of papers already published using this approach (1–5). Nonetheless in the future it may still be more convenient or proportionate to execute analyses only in OpenSAFELY-TPP; therefore the high degree of representativeness demonstrated in this paper provides strong reassurance that such analyses present no interpretive or generalisability challenges.

## Conclusions

Despite regional variations, data from OpenSAFELY-TPP is largely representative of the general population of England in terms of IMD, age, sex, ethnicity and causes of death. Following the use of OpenSAFELY-TPP data for a large number of COVID-19 studies since March 2020, it is reassuring to find that the overall representativeness of the population in the datasets is high.

## Data Availability

Access to the underlying identifiable and potentially re-identifiable pseudonymised electronic health record data is tightly governed by various legislative and regulatory frameworks, and restricted by best practice. The data in OpenSAFELY is drawn from General Practice data across England where TPP is the Data Processor. TPP developers (CB, JC, JP, FH, and SH) initiate an automated process to create pseudonymised records in the core OpenSAFELY database, which are copies of key structured data tables in the identifiable records. These are linked onto key external data resources that have also been pseudonymised via SHA-512 one-way hashing of NHS numbers using a shared salt. DataLab developers and PIs (BG, LS, CEM, SB, AJW, KW, WJH, HJC, DE, PI, SD, GH, BBC, RMS, ID, KB, SE, EJW and CTR) holding contracts with NHS England have access to the OpenSAFELY pseudonymised data tables as needed to develop the OpenSAFELY tools. These tools in turn enable researchers with OpenSAFELY Data Access Agreements to write and execute code for data management and data analysis without direct access to the underlying raw pseudonymised patient data, and to review the outputs of this code.

## Abbreviations

EHR: Electronic Health Record
ICD: International Classification of Diseases
IMD: Index of Multiple Deprivation
NUTS: Nomenclature of Territorial Units for Statistics
ONS: Office for National Statistics
SUS: Secondary Uses Service

## Administrative

## Acknowledgements

We are very grateful for all the support received from the TPP Technical Operations team throughout this work, and for generous assistance from the information governance and database teams at NHS England / NHSX.

## Information governance and ethical approval

NHS England is the data controller; TPP is the data processor; and the researchers on OpenSAFELY are acting with the approval of NHS England. This implementation of OpenSAFELY is hosted within the TPP environment which is accredited to the ISO 27001 information security standard and is NHS IG Toolkit compliant;(22,23) patient data has been pseudonymised for analysis and linkage using industry standard cryptographic hashing techniques; all pseudonymised datasets transmitted for linkage onto OpenSAFELY are encrypted; access to the platform is via a virtual private network (VPN) connection, restricted to a small group of researchers; the researchers hold contracts with NHS England and only access the platform to initiate database queries and statistical models; all database activity is logged; only aggregate statistical outputs leave the platform environment following best practice for anonymisation of results such as statistical disclosure control for low cell counts.(24) The OpenSAFELY research platform adheres to the obligations of the UK General Data Protection Regulation (GDPR) and the Data Protection Act 2018. In March 2020, the Secretary of State for Health and Social Care used powers under the UK Health Service (Control of Patient Information) Regulations 2002 (COPI) to require organisations to process confidential patient information for the purposes of protecting public health, providing healthcare services to the public and monitoring and managing the COVID-19 outbreak and incidents of exposure; this sets aside the requirement for patient consent.(25) Taken together, these provide the legal bases to link patient datasets on the OpenSAFELY platform. GP practices, from which the primary care data are obtained, are required to share relevant health information to support the public health response to the pandemic, and have been informed of the OpenSAFELY analytics platform.

This study was approved by the Health Research Authority (REC reference 20/LO/0651) and by the LSHTM Ethics Board (reference 21863).

## Data access and verification

Access to the underlying identifiable and potentially re-identifiable pseudonymised electronic health record data is tightly governed by various legislative and regulatory frameworks, and restricted by best practice. The data in OpenSAFELY is drawn from General Practice data across England where TPP is the Data Processor. TPP developers (CB, JC, JP, FH, and SH) initiate an automated process to create pseudonymised records in the core OpenSAFELY database, which are copies of key structured data tables in the identifiable records. These are linked onto key external data resources that have also been pseudonymised via SHA-512 one-way hashing of NHS numbers using a shared salt. DataLab developers and PIs (BG, LS, CEM, SB, AJW, KW, WJH, HJC, DE, PI, SD, GH, BBC, RMS, ID, KB, SE, EJW and CTR) holding contracts with NHS England have access to the OpenSAFELY pseudonymised data tables as needed to develop the OpenSAFELY tools. These tools in turn enable researchers with OpenSAFELY Data Access Agreements to write and execute code for data management and data analysis without direct access to the underlying raw pseudonymised patient data, and to review the outputs of this code. All code for the full data management pipeline—from raw data to completed results for this analysis—and for the OpenSAFELY platform as a whole is available for review at github.com/OpenSAFELY.

The data management and analysis code for this paper was led by CDA and contributed to by WJH and AJW.

## Guarantor

BG is guarantor.

## Contributorship

*All authors contributed to and approved the final manuscript*.

